# Assessment of Joint and Interactive Effects of Multimorbidity and Chronic Pain on ADRD Risk in the Elder Population

**DOI:** 10.1101/2022.08.02.22278338

**Authors:** Sumaira Khalid, Kim E. Innes, Amna Umer, Christa Lilly, Diane Gross, Usha Sambamoorthi

## Abstract

**Objective:** Multimorbidity and non-cancer chronic pain conditions (NCPC) are independently linked to elevated risk for cognitive impairment and incident Alzheimer’s Disease and Related Dementias (ADRD)-both - We present the study of potential joint and interactive effects of these conditions on the risk of incident ADRD in older population.

**Methods:** This retrospective-cohort study drew baseline and 2-year follow-up data from linked Medicare claims and Medicare Current Beneficiary Survey (MCBS). Baseline multimorbidity and NCPC were ascertained using claims data. ADRD was ascertained at baseline and follow-up.

**Results:** NCPC accompanied by multimorbidity (vs. absence of NCPC or multimorbidity) had a significant and upward association with incident ADRD (adjusted odds ratio (AOR): 1.72, 95% CI 1.38, 2.13, *p<*0.0001). Secondary analysis by number of comorbid conditions suggested that the joint effects of NCPC and multimorbidity on ADRD risk may increase with rising number contributing chronic conditions. Interaction analyses indicated significantly elevated excess risk for incident ADRD.

## 1. Introduction

A dramatic rise in prevalence of Alzheimer’s disease and related dementias (ADRD), both worldwide and in the United States (U.S.), is projected in the next decade owing in part to increasing longevity and growing number of older adults (“Alzheimer’s Association Facts and Figures,” 2021). Paralleling the increase in ADRD prevalence is a corresponding escalation in disability, dependence, and institutionalization rates accompanied by overwhelming healthcare costs (Nichols et al., 2019; Takizawa, Thompson, van Walsem, Faure, & Maier, 2015). To effectively target at risk populations for preventing or slowing conversion to ADRD, understanding the interaction between multiple factors affecting ADRD risk, especially those amenable to change, is paramount. Among such modifiable risk factors, certain chronic conditions including diabetes, heart and cerebrovascular disease, hypertension, obesity, serious respiratory disorders (Baumgart et al., 2015; Cooper, Sommerlad, Lyketsos, & Livingston, 2015) As well as depression(Ownby, Crocco, Acevedo, John, & Loewenstein, 2006), anxiety (Gulpers et al., 2016) and chronic disorders of sleep (Bubu et al., 2016) have also been associated to risk for cognitive decline and ADRD.

Multimorbidity is on the rise globally as well as in the U.S. Multimorbidity, defined as the concurrent presence of 2 or more chronic conditions (Johnston, Crilly, Black, Prescott, & Mercer, 2018), has traditionally been overlooked in ADRD epidemiology where individual risk factors were included and adjusted for in the epidemiologic analyses. More recent developments suggest that the study of interaction between chronic comorbid conditions is as important as their individual effects on the risk for ADRD. In addition, underreported chronic conditions often get overlooked while studying multimorbidity. For example, non-cancer chronic pain, a highly frequent yet understudied condition in older population. Accumulating evidence suggests that both multimorbidity and certain non-cancer chronic pain conditions (NCPC), for example headache, fibromyalgia, knee and joint pain, and neuropathic pain may likewise significantly increase risk for cognitive decline and new-onset dementia (Innes & Sambamoorthi, 2020; Wei, Levine, Zahodne, Kabeto, & Langa, 2020). Like that of ADRD, the prevalence of both multimorbidity and NCPCs increases steeply with advancing age, and there is also growing evidence that multimorbidity is strongly and reciprocally associated with chronic pain and related functional impairment, psychological distress, and other adverse sequalae (Nakad et al., 2020), factors also linked to poor cognitive outcomes (Lacreuse, Raz, Schmidtke, Hopkins, & Herndon, 2020). Extant evidence, although limited, suggests that multimorbidity can exacerbate chronic pain and vice versa (Nakad et al., 2020; Scherer et al., 2016), contributing to a vicious cycle of deteriorating health and function which may, in turn, further increase the risk for accelerated cognitive decline, cognitive impairment, and incident ADRD.

However, although there is some documented evidence of independent associations of NCPCs (Innes & Sambamoorthi, 2020) and multimorbidity (Wei et al., 2020) to ADRD risk, and growing evidence presents clues of an important and potentially synergistic role of both multimorbidity and chronic pain in the development of cognitive decline and ADRD in older adults (Jellinger & Attems, 2015; Nakad et al., 2020), the potential joint and interactive effect of chronic pain and multimorbidity involving other chronic health conditions on risk for incident ADRD remains unexplored. While chronic pain, a chronic health condition itself, is a leading cause of years lived with disability in the population, it suffers an underrepresentation in the published multimorbidity literature. In this study, we used linked data from a large, nationally representative sample of older United States Medicare beneficiaries to investigate the potential joint and interactive effects of multimorbidity from other chronic health conditions and NCPCs on incident ADRD. We hypothesized that co-occurring NCPCs and multimorbidity would amplify the effects of both conditions on ADRD risk, and that the joint effects of NCPC and multimorbidity would be more pronounced with a higher number of chronic conditions contributing to multimorbidity.

## 2. Methods

### 2.1. Study Design and Data Source

Multiple Medicare Current Beneficiary Survey (MCBS) cohorts from several years (2001-2013) were pooled to generate a large study sample with sufficient study power to evaluate the joint effect of baseline multimorbidity and NCPCs on risk for incident ADRD. We employed a retrospective cohort study design, and our study cohorts were drawn from a nationally representative sample of Medicare fee-for-service beneficiaries 65+ years of age who completed the Medicare Current Beneficiary Survey (MCBS). MCBS uses a complex stratified, three-stage probability sampling design for participant recruitment, and collects both cross-sectional and longitudinal data on health status, health services utilization, prescriptions, and healthcare costs using a series of survey interviews and administrative records; comprehensive explanation of procedures can be found in previous publications (MCBS; Mues et al., 2017). Our study specifically used MCBS Cost and Use files linked with Medicare fee-for-service claims to ascertain demographics, access to care, lifestyle factors, medical conditions and medication use.

### 2.2. Eligibility Criteria and Analytic sample

The study analytic sample comprised non-institutionalized MCBS participants aged 65+ years at baseline, alive at the end of follow-up (defined as the time of incident ADRD diagnosis or end of the 3-year study period) and enrolled in Medicare fee-for-service (FFS) throughout the 3-year study period or until incident ADRD diagnosis. Participants with diagnosed ADRD at baseline were excluded. Applying a priori exclusion criteria yielded a final analytic sample size of 16,934 adults.

### 2.3. Measures

#### Incident Alzheimer’s Disease and Related Dementia (ADRD)

We ascertained ADRD status at baseline year and during the study follow-up years 2 and 3, using FFS claims for inpatient (IP), skilled nursing facility (SNF), outpatient (OT), home health agency (HHA), and physician office (PO) visits for years 2001–2013 as well as MCBS self-reported Health Status and Functioning files. To this end, we used a validated algorithm (Centers for Medicare and Medicaid Services (CMS) Chronic Condition Algorithms) (“Chronic Conditions Data Warehouse: Your source for national CMS Medicare and Medicaid research data,” Access Yr 2020) of at least one fee-for-service claim with any of the following International Classification of Diseases, ninth Edition, clinical modification (ICD-9-CM) diagnostic codes: 331.0, 331.11, 331.19, 331.2, 331.7, 290.0, 290.10, 290.11, 290.12, 290.13, 290.20, 290.21, 290.3, 290.40, 290.41, 290.42, 290.43, 294.0, 294.10, 294.11, 294.20, 294.21, 294.8, and 797, or an affirmative response to the self-reported Health Status question “Has a doctor ever told you that you had Alzheimer’s?” (Lin et al., 2010). Using a combination of claims and survey data to ascertain ADRD has been recommended by MCBS investigators to increase capture of ADRD, and has been shown to yield results similar to those of expert in-person-assessments (Rose et al., 2016).

#### Presence of Non-Cancer Chronic Pain Conditions (NCPCs)

FFS claims were used to ascertain NCPC status at baseline as either two outpatient claims (90 days apart) or one inpatient claim using ICD-9-CM codes as recommended by CMS (“Chronic Conditions Data Warehouse: Your source for national CMS Medicare and Medicaid research data,” Access Yr 2020) and consistent with prior studies of NCPC (Sullivan, Turner, & Romano, 1991). Any NCPC was assessed as a binary variable (yes/no) during baseline, including five common non-cancer pain conditions: back or neck pain, headache, joint pain, neuropathic pain, and osteoarthritis.

#### Presence of Multimorbidty

Multimorbidity was ascertained using FFS claims during the baseline year using ICD-9-CM (“Chronic Conditions Data Warehouse: Your source for national CMS Medicare and Medicaid research data,” Access Yr 2020). The presence of multimorbidity was defined as having 2 or more co-existing chronic physical health conditions (excluding NCPCs) in addition to the index disease (ADRD) (Buntinx & Knottnerus, 1996). The construction of multimorbidity variable was guided by the conceptual framework on multiple chronic conditions detailed by Richard Goodman et al. (Goodman et al., 2016), which provided standardized definitions and identification approaches to improve the understanding and surveillance of individual as well as multiple co-occurring conditions (multimorbidity) implicated in high disease burden and escalating healthcare costs in the United States. Analytical variables for multimorbidity were constructed using 1) an aggregate binary variable (yes/no) defined using a comprehensive set of diagnosed chronic conditions, other than NCPCs, at baseline, including: asthma, cardiac arrythmias, cerebrovascular disease, coronary artery disease, congestive heart failure, chronic obstructive pulmonary disease, cancer, diabetes, hypercholesterolemia, and hypertension; 2) to study the effects of number of chronic conditions comprising multimorbidity, a secondary variable was created with 3 categories: multimorbidity with 2-3 chronic conditions, multimorbidity with ≥ 4 chronic conditions, and no multimorbidity (reference category, < 2 chronic conditions).

#### Covariates

Factors widely reported or suspected to be associated with ADRD risk, chronic pain, and/or multimorbidity, were accounted for, using a priori inclusion criteria in our multivariable models. These included biological factors (age, sex and race/ethnicity (Non-Hispanic White, Non-Hispanic Black, Hispanic, and other)), demographic characteristics (marital status, educational level, family income, health insurance status), lifestyle factors (smoking status, body mass index), History of other chronic conditions at baseline not included in the multimorbidity variable, such as traumatic brain injury (TBI) and specific auto-immune conditions, rheumatoid arthritis (RA), and systemic lupus erythematosus (SLE). Common medications that have been linked to ADRD risk and/or used in chronic pain management were also evaluated as covariates. These included non-steroidal anti-inflammatory drugs (NSAIDS) (Etminan, Gill, & Samii, 2003) and opioid analgesics (Pask, Dell’Olio, Murtagh, & Boland, 2019). Since mood and sleep impairment have been strongly and reciprocally associated with both multimorbidity (Goodman et al., 2016; Helbig et al., 2017; Read, Sharpe, Modini, & Dear, 2017) and chronic pain/NCPCs (McWilliams, Goodwin, & Cox, 2004; Menefee et al., 2000; Zis et al., 2017), and may in part mediate the observed joint effects on incident ADRD risk, these conditions were evaluated separately as diagnosed depression, anxiety and insomnia-related sleep disorders, ascertained at baseline using ICD-9-CM codes (“Chronic Conditions Data Warehouse: Your source for national CMS Medicare and Medicaid research data,” Access Yr 2020).

### 2.4. Statistical Analysis

Differences in baseline characteristics by multimorbidity were assessed using Chi-square tests. Logistic regressions were used to assess the unadjusted and adjusted individual and joint associations of NCPCs and multimorbidity to incident ADRD. In the multivariable logistic regressions, we carried out incremental block-wise adjustment for demographic and socioeconomic factors, lifestyle factors, certain other chronic health conditions, and analgesics use. Effect of mood and sleep disorders on the adjusted joint associations of NCPC and multimorbidity to ADRD risk was assessed separately.

Investigation of joint and interactive effects in this study was guided by the recommendations for reporting effect modification and interactions in epidemiologic research by Knol and VanderWeele (Knol & VanderWeele, 2012). Specifically, the recommendations involve reporting of 1) individual associations (odds ratios (OR)) of both the key independent variables; 2) joint effects (OR) of the key independent variables on the association with the outcome; 3) the relative excess risk due to interaction (RERI) for reporting excess risk on additive scale due to the presence of another key variable, as well as the risk due to interaction on multiplicative scale. To investigate the added risk of NCPC specifically for each level of multimorbidity, i.e., multimorbidity with 2-3 chronic conditions and with ≥4 chronic conditions, we performed regression analyses with contrast using respective reference categories for each contrast.

For assessment of potential interactions in this study, we calculated individual (OR_NCPC, no Multimorbidity_, OR_Multimorbidity, no NCPC_) and joint (OR_NCPC, Multimorbidity_) effects, using the same reference category (no multimorbidity and no NCPC). Additive risk due to interaction was calculated using RERI, whereas separate logistic regression contrast models for 2-3 chronic conditions and ≥ 4 chronic conditions were used to compare the increment in the risk in the presence of NCPC. Multiplicative interaction was evaluated by employing an interaction term in the logistic regression models. For an adequate comparison, each joint and interactive effect was assessed across all adjusted analyses.

All analyses were weighted and used MCBS 3-year backward longitudinal weights. All analyses were performed using SAS survey procedure (SAS version 9.4, SAS Institute, Inc.) and R Studio software (version 1.0.136, RStudio, Inc.).

## 3. Results

Our analytic sample comprised 16,934 eligible participants. Table 1 summarizes the study characteristics distributed by the presence or absence of multimorbidity. The majority of our analytic sample with multimorbidity were female (58%), white (84%), married (52%), lived in metropolitan areas (73%), had private insurance (80%), and reported a family income at or above 200% of the FPL (51%). Participants with multimorbidity had higher use of opioid analgesics (22% vs. 12%) and NSAIDs (21% vs. 13%). Overall baseline prevalence of NCPCs in the multimorbidity group was 68% (weighted). The percentage of participants with baseline NCPCs varied significantly by number of co-occurring chronic conditions ((76% vs. 62% vs. 39%, for ≥4 vs. 2-3 vs. no multimorbidity, respectively, p<0.0001) (data not shown). The proportion of participants with diagnosed depression, anxiety, or sleep disorders at baseline also rose with increasing number of comorbid conditions (p’s <0.0001). Participants with ≥4 co-occurring chronic conditions (vs. 2-3 and no multimorbidity) were also more likely to be older (80+), on Medicaid insurance, and to report lower family income.

**Table 1.**
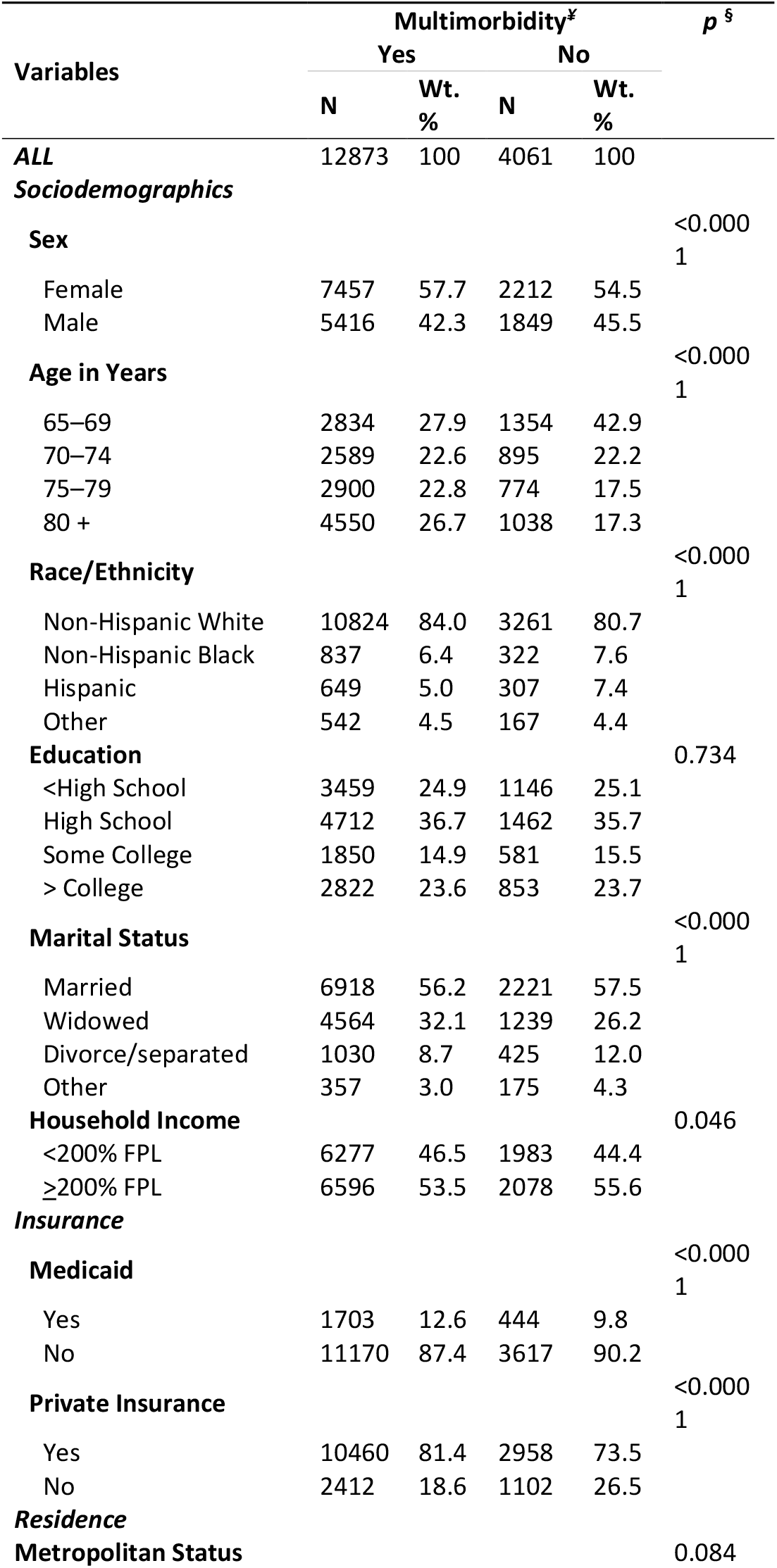
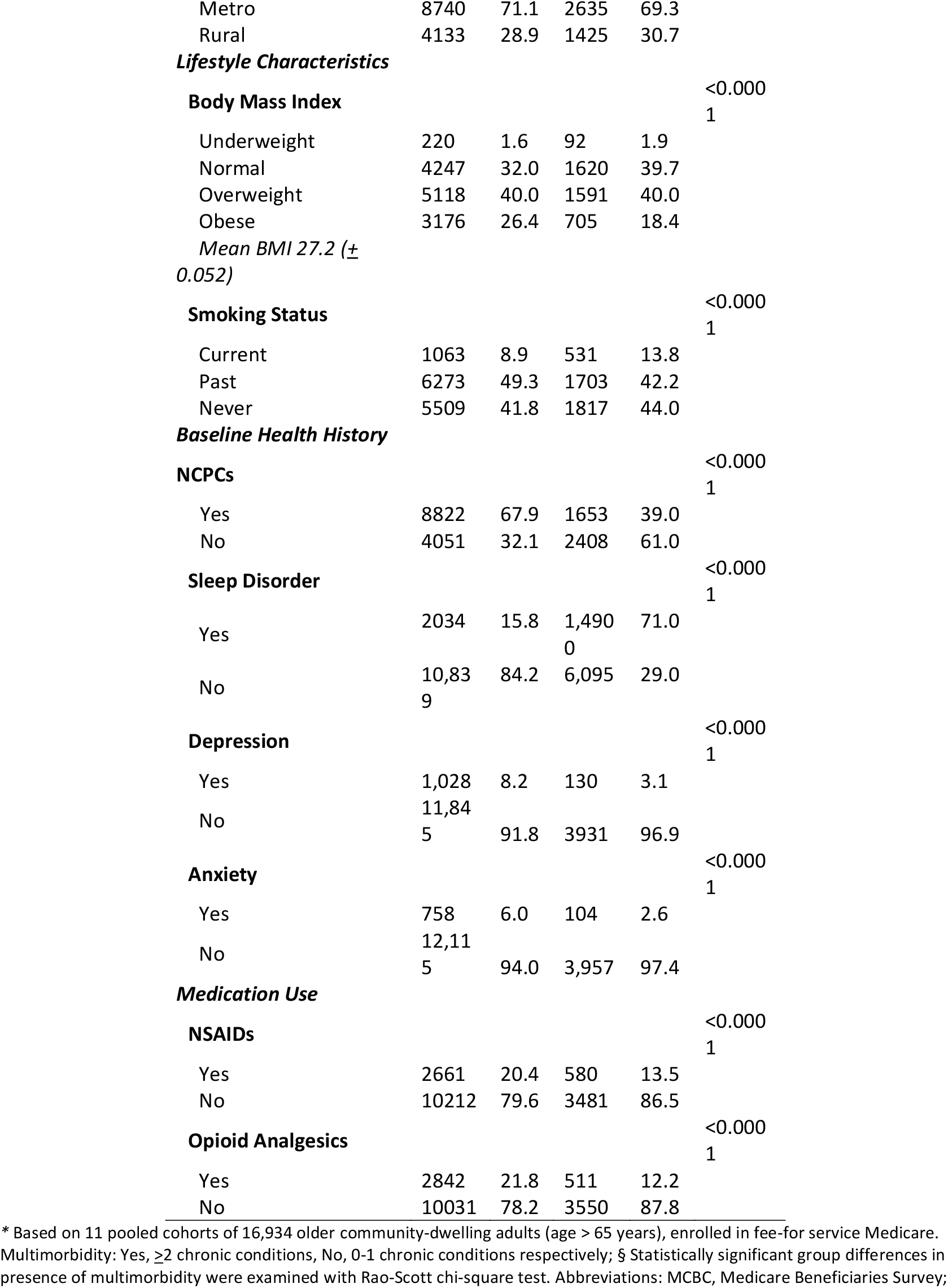
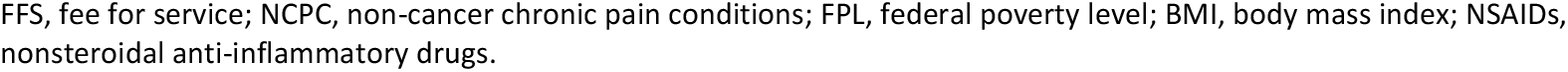
Baseline characteristics of study participants* stratified by multimorbidity, analyzed using linked MCBS and FFS claims data, 2001–2013.

At the end of study follow-up period, 68 for every 1000 participants were diagnosed with incident ADRD (N=1149). Incident ADRD rates were significantly higher in those with NCPC vs. without baseline NCPCs (7.3% vs 4.7%) and multimorbidity (6.3% vs 4.0%) (*p*’s < 0.0001). Incident ADRD rates were also higher in women, non-Hispanic blacks, widowed, poorly educated, and participants <200% FPL, Medicaid insurance and those who lacked private insurance (*p*’s < 0.0001). Those with a history of TBI, as well as those who reported opioid analgesics use had a higher incidence rate compared to those without (*p*’s<0.0001). Similarly, those who were diagnosed with sleep, depression or anxiety disorders at baseline included a significantly higher proportion of incident ADRD cases (*p*’s < 0.0001) (see supplementary table).

### 3.1. Joint and interactive effects of multimorbidity and NCPC on risk for incident ADRD

Table 2 illustrates the joint and interactive effects of multimorbidity and NCPC on ADRD risk. Compared to participants with no NCPC and multimorbidity at baseline (reference category), those with concomitant NCPC and multimorbidity were significantly more likely to be diagnosed with incident ADRD at follow-up (unadjusted odds ratios (ORs) = 2.07, 95% confidence interval (CI) 1.67, 2.55, p<0.0001). This association remained robust even after adjustment for covariates (adjusted ORs (AOR’s) = 1.72, CI= 1.38, 2.13, *p* < 0.0001). Likewise, risk estimates for multimorbidity and NCPC alone were only modestly reduced in the fully adjusted analyses, although only multimorbidity alone remained significantly associated with risk for incident ADRD (AOR’s for multimorbidity and presence of NCPCs alone, respectively, =1.31 (1.07,1.60) and 1.25 (0.87,1.78)). Further analysis by number of co-occurring chronic conditions yielded similar findings (table-2b); the combined presence of NCPC and multimorbidity remained strongly and positively associated with incident ADRD in the fully adjusted analyses, with risk for ADRD highest in those with the greatest number of chronic conditions (AOR’s for NCPC and ≥4 chronic conditions vs no NCPC and < 2 chronic conditions =1.88 (1.68, 2.40), p<0.0001). Contrast analyses, using 2-3 and ≥ 4 chronic conditions without NCPC as reference categories, indicated significantly elevated risk for ADRD in those with co-occurring NCPC (AORs for NCPC accompanied by 2-3 and ≥ 4 chronic conditions, respectively = 1.32 (1.08, 1.62, *p=*0.0076); 1.23 (1.01, 1.51), *p=*0.0499).

**Table 2a.**
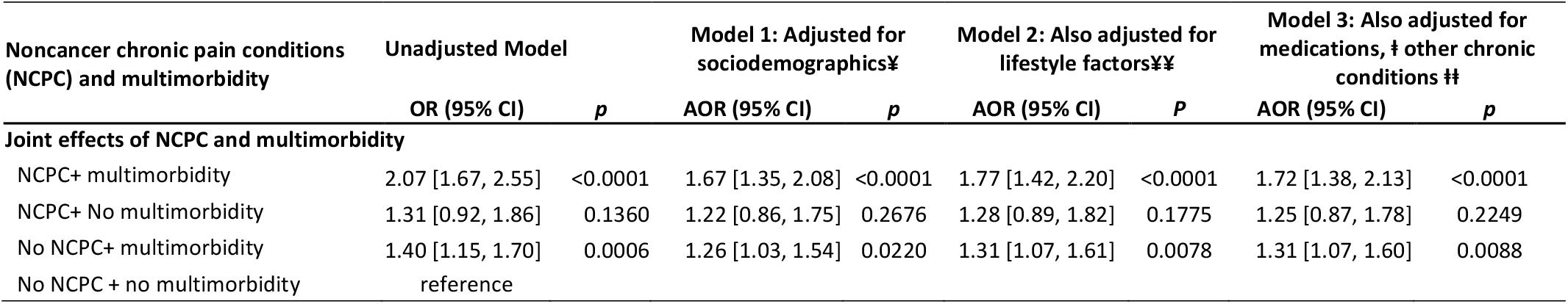
Effects of baseline non-cancer chronic pain conditions (NCPC) and multimorbidity, alone and in combination, on incident Alzheimer’s disease and related dementias (ADRD): analysis using linked Medicare Current Beneficiary Survey and Medicare claims data*

| Noncancer chronic pain conditions (NCPC) and multimorbidity | Unadjusted Model |  | Model 1: Adjusted for sociodemographics¥ |  | Model 2: Also adjusted for lifestyle factors¥¥ |  | Model 3: Also adjusted for medications, † other chronic conditions ‡ |  |
| --- | --- | --- | --- | --- | --- | --- | --- | --- |
|  | OR (95% CI) | *p* | AOR (95% CI) | *p* | AOR (95% CI) | *P* | AOR (95% CI) | *p* |
| **Joint effects of NCPC and multimorbidity** |  |  |  |  |  |  |  |  |
| NCPC+ multimorbidity | 2.07 [1.67, 2.55] | <0.0001 | 1.67 [1.35, 2.08] | <0.0001 | 1.77 [1.42, 2.20] | <0.0001 | 1.72 [1.38, 2.13] | <0.0001 |
| NCPC+ No multimorbidity | 1.31 [0.92, 1.86] | 0.1360 | 1.22 [0.86, 1.75] | 0.2676 | 1.28 [0.89, 1.82] | 0.1775 | 1.25 [0.87, 1.78] | 0.2249 |
| No NCPC+ multimorbidity | 1.40 [1.15, 1.70] | 0.0006 | 1.26 [1.03, 1.54] | 0.0220 | 1.31 [1.07, 1.61] | 0.0078 | 1.31 [1.07, 1.60] | 0.0088 |
| No NCPC + no multimorbidity | reference |  |  |  |  |  |  |  |

**Table 2b.** Interactive effects of baseline non-cancer chronic pain conditions (NCPC) and multimorbidity on incident Alzheimer’s disease and related dementias (ADRD): analysis from linked Medicare Current Beneficiary Survey and Medicare claims data*

| Noncancer chronic pain conditions (NCPC) and number of chronic conditions | Unadjusted Model |  | Model 1: Adjusted for sociodemographics¥ |  | Model 2: Also adjusted for lifestyle factors¥¥ |  | Model 3: Also adjusted for medications, ‡ other chronic conditions ‡‡ |  |
| --- | --- | --- | --- | --- | --- | --- | --- | --- |
|  | OR (95% CI) | p | AOR (95% CI) | p | AOR (95% CI) | P | AOR (95% CI) | p |
| **Joint effects of NCPC and multimorbidity by number of co-occurring chronic conditions** |  |  |  |  |  |  |  |  |
| NCPC + ≥4 conditions | 2.37 [1.87,3.00] | <0.0001 | 1.83[1.63,2.33] | <0.0001 | 1.94[1.62,2.47] | <0.0001 | 1.88[1.68,2.40] | <0.0001 |
| NCPC + 2-3 conditions | 1.79 [1.43,2.23] | <0.0001 | 1.53[1.22,1.92] | 0.0003 | 1.62[1.28,1.92] | <0.0001 | 1.57[1.25,1.78] | <0.0001 |
| No NCPC + ≥4 conditions | 1.75 [1.40,2.18] | <0.0001 | 1.48[1.17,1.87] | 0.0011 | 1.55[1.22,1.97] | 0.0003 | 1.53[1.21,1.94] | 0.0004 |
| No NCPC + 2-3 conditions | 1.22 [0.99,1.52] | 0.0624 | 1.15[0.93,1.42] | 0.2078 | 1.20[0.96,1.49] | 0.1040 | 1.19[0.96,1.48] | 0.1087 |
| No NCPC + <2 conditions | reference |  |  |  |  |  |  |  |
| **Additive risk due to interaction of NCPC and number of chronic conditions\*\*** |  |  |  |  |  |  |  |  |
| NCPC + 2-3 conditions | 1.46 [1.19,1.78] | 0.0002 | 1.33[1.09,1.63] | 0.0054 | 1.35[1.10,1.65] | 0.0036 | 1.32[1.08, 1.62] | 0.0076 |
| No NCPC + 2-3 conditions | reference |  |  |  |  |  |  |  |
| NCPC + ≥4 conditions | 1.36 [1.11,1.65] | 0.0027 | 1.23[1.01,1.51] | 0.0437 | 1.25[1.22,1.54] | 0.0277 | 1.23[1.01, 1.51] | 0.0499 |
| No NCPC + ≥4 conditions | reference |  |  |  |  |  |  |  |

\*Study sample comprised 11 pooled cohorts (2001–2013) of 16,934 older community-dwelling adults aged 65+ years and enrolled in fee-for-service Medicare. Odds ratios (OR) and adjusted odds ratios (AOR) with 95% confidence intervals (CI) were calculated using separate logistic regression models. ¥ Including socio-demographics: sex, age, race/ethnicity, education, income, Medicaid insurance, private insurance, marital status, region. ¥¥ Also including lifestyle factors: smoking status, body mass index; ‡ Also including history of traumatic brain injury, use of nonsteroidal anti-inflammatory drugs and opioid analgesics, \*\*contrast using logistic regression models; p-value from multiplicative interactions from fully adjusted logistic regression p=0.773 indicates absence of multiplicative interaction

Inclusion of potential mediators, i.e., sleep disorders, depression, and anxiety, in the fully adjusted models attenuated but did not eliminate the joint effects of NCPCs and multimorbidity on ADRD risk (AOR’s for combined presence of NCPC and multimorbidity and of NCPC and 2-3 and ≥4 conditions, respectively = 1.45 (1.16, 1.81), *p* < 0.001; 1.40 (1.11, 1.76), p<0.005; and 1.53 (1.19, 1.97), p<0.0001).

Additional analysis assessing relative excess risk for incident ADRD due to interaction between multimorbidity and NCPC (RERI) indicated that the additive interaction of NCPC with multimorbidity accounted for a relative excess risk of 36% (29.3, 42.6) (Table-2c). As shown in table-2c, the additive interaction of NCPC with 2-3 and ≥4 co-occurring chronic conditions accounted for a relative excess risk of 31% (23.5, 38.5) and 26% (18.7, 33.2), respectively, after accounting for the individual effects of these factors. These interactive effects were attenuated but remained statistically significant after adjustment for demographics, socioeconomic characteristics, other chronic condition, and analgesic use (RERI (%) for NCPC with multimorbidity, 2-3, and ≥4 chronic conditions, respectively = 16% (10.1, 21.8); 10% (3.9, 16.0); and 13% (6.6, 19.3)). In contrast, we found no evidence for a multiplicative interaction of NCPCs and either 2-3 conditions or ≥4 chronic conditions in the fully adjusted models (p’s>0.1)).

**Table 2c.**
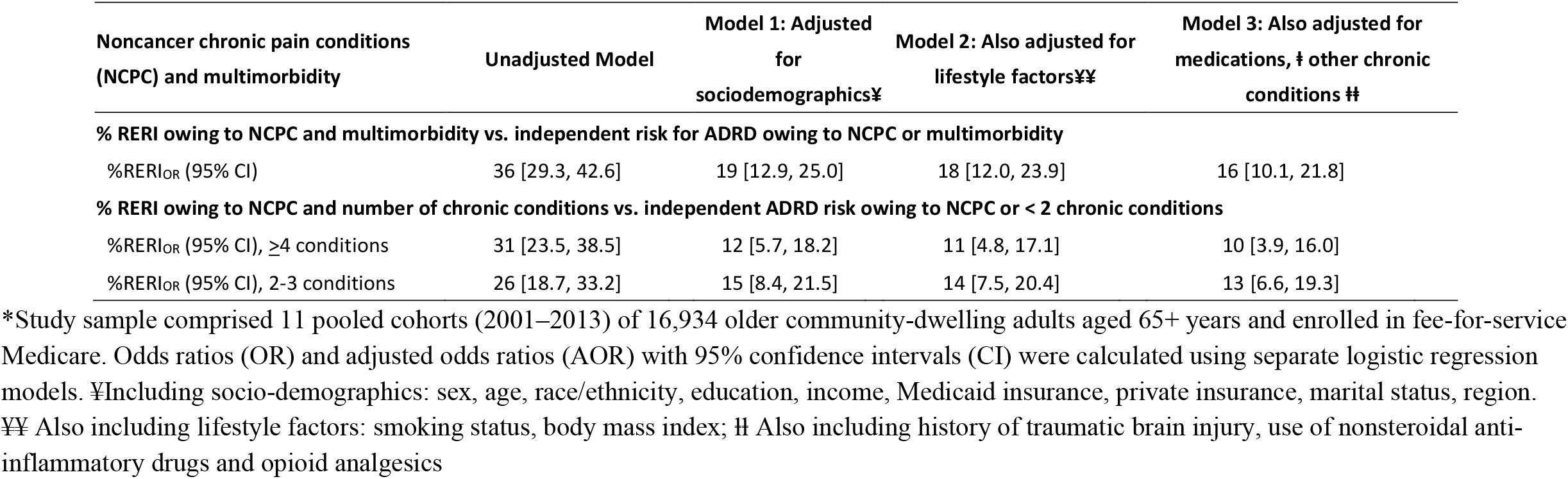
Relative excess risk due to interaction (RERI) of baseline non-cancer chronic pain conditions (NCPC) and multimorbidity on incident Alzheimer’s disease and related dementias (ADRD): analysis from linked Medicare Current Beneficiary Survey and Medicare claims data*

| Noncancer chronic pain conditions (NCPC) and multimorbidity | Unadjusted Model | Model 1: Adjusted for sociodemographics¥ | Model 2: Also adjusted for lifestyle factors¥¥ | Model 3: Also adjusted for medications, ‡ other chronic conditions ‡‡ |
| --- | --- | --- | --- | --- |
| **% RERI owing to NCPC and multimorbidity vs. independent risk for ADRD owing to NCPC or multimorbidity** |  |  |  |  |
| %RERI<sub>OR</sub> (95% CI) | 36 [29.3, 42.6] | 19 [12.9, 25.0] | 18 [12.0, 23.9] | 16 [10.1, 21.8] |
| **% RERI owing to NCPC and number of chronic conditions vs. independent ADRD risk owing to NCPC or < 2 chronic conditions** |  |  |  |  |
| %RERI<sub>OR</sub> (95% CI), ≥4 conditions | 31 [23.5, 38.5] | 12 [5.7, 18.2] | 11 [4.8, 17.1] | 10 [3.9, 16.0] |
| %RERI<sub>OR</sub> (95% CI), 2-3 conditions | 26 [18.7, 33.2] | 15 [8.4, 21.5] | 14 [7.5, 20.4] | 13 [6.6, 19.3] |

\*Study sample comprised 11 pooled cohorts (2001–2013) of 16,934 older community-dwelling adults aged 65+ years and enrolled in fee-for-service Medicare. Odds ratios (OR) and adjusted odds ratios (AOR) with 95% confidence intervals (CI) were calculated using separate logistic regression models. ¥Including socio-demographics: sex, age, race/ethnicity, education, income, Medicaid insurance, private insurance, marital status, region. ¥¥ Also including lifestyle factors: smoking status, body mass index; ‡ Also including history of traumatic brain injury, use of nonsteroidal anti-inflammatory drugs and opioid analgesics

## 4. Discussion

To our knowledge, this investigation is the first large retrospective cohort study to assess the joint and interactive effects of NCPC and multimorbidity, highly prevalent conditions in older adults, on the risk for incident ADRD, and to explore the potential synergistic effects of NCPC and chronic condition burden on incident ADRD risk in community-dwelling older adults. We found that participants with both NCPCs and multimorbidity at baseline were at significantly elevated risk for incident ADRD after adjustment for multiple potential confounders, with risk estimates substantially exceeding those for either NCPC or multimorbidity alone. Further analysis regarding the joint effects of NCPCs and multimorbidity by number of co-occurring chronic conditions showed similar results i.e., NCPC in the presence of both low (2-3 conditions) and high (≥4) chronic condition burden was significantly and positively associated with risk for incident ADRD, with risk estimates particularly pronounced in those with NCPC and ≥ 4 chronic conditions. Additional analyses regarding the interactive effects of NCPC’s and multimorbidity indicated that the additive interaction of these conditions accounted for a significant positive excess risk for incident ADRD after adjustment for potential confounders. Excess risk measure (RERI) yielded findings consistent with the primary analyses across all adjusted models; risk for ADRD remained significantly higher in the presence of co-occurring NCPC’s and multimorbidity compared to the presence of either condition alone. In agreement with our primary hypothesis, these findings suggest that multimorbidity may amplify the effects of NCPC on risk for ADRD and vice versa. Our findings also suggest that the joint effects of NCPC and multimorbidity on ADRD risk may increase with rising number of contributing chronic conditions.

A number of large, population-based studies in the U.S. (Khalid, Sambamoorthi, & Innes, 2020; Whitlock et al., 2017), European (Hagen et al., 2014; Røttereng et al., 2015), Canadian (Morton, St. John, & Tyas, 2019), and East Asian adults (Tzeng et al., 2018; Yamada et al., 2019) have reported significant, positive associations of certain common chronic pain conditions, including headache, osteoarthritis and knee pain and fibromyalgia, to subsequent risk for ADRD after adjustment for demographics, certain chronic conditions, and other potential confounders. Likewise, a growing number of observational studies suggest that multimorbidity is an important predictor of cognitive decline (Vassilaki et al., 2015) and late-onset ADRD (Duthie, Chew, & Soiza, 2011; Haaksma et al., 2017; Jellinger & Attems, 2015). For example, a prospective cohort study in cognitively normal community elders in Olmsted county, Minnesota, demonstrated heightened risk for developing mild cognitive impairment and dementia in individuals with both ≥ 2 chronic conditions and ≥ 4 chronic conditions (Vassilaki et al., 2015). However, although prior investigations have made efforts to adjust for the potential confounding influence of individual chronic health conditions, the potential joint and interactive effect of other comorbid chronic conditions with NCPC on the association between NCPCs and incident ADRD has remained unexplored.

Prevalence of multimorbidity, like that of NCPC (Dahlhamer et al., 2018), is rising globally as well as in the U.S. For example, in the U.S. almost three-quarters of adults 65 years of age and older have at least 2 concomitant chronic conditions, and over one-third have at least 4 chronic conditions (Goodman et al., 2016). Moreover, evidence from observational studies suggests that multimorbidity not only increases in prevalence with advancing age (Marengoni et al., 2011), but is strongly and bidirectionally associated with chronic pain (Nakad et al., 2020; Scherer et al., 2016). Recent studies also indicate that chronic pain and multimorbidity share many risk factors and sequalae (Prados-Torres, Calderón-Larranaga, Hancco-Saavedra, Poblador-Plou, & van den Akker, 2014). For example, a recent review summarizing the relationship and outcomes of multimorbidity and chronic pain in older adults, documented deterioration in functional and psychological faculties, poor quality of life, and impaired sleep among other consequences of pain and multimorbidity co-occurrence (Nakad et al., 2020). Furthermore, the combined presence of multimorbidity and NCPC can exacerbate functional impairment and sedentarism (Calderón-Larrañaga et al., 2019), factors also linked to chronic pain (Geneen et al., 2017) and to increased risk for ADRD (Baumgart et al., 2015).

Although the mechanisms underlying the apparent synergistic effects of multimorbidity and NCPCs on risk for incident ADRD are incompletely understood, multimorbidity may contribute to increased ADRD risk in older adults, both independently and in concert with NCPCs, via several pathways. Recent multimorbidity research has provided support for potential disease-disease interactions among certain comorbid chronic conditions, including complex pathophysiologic interactions, that may contribute to the development of neurodegenerative disorders such as ADRD (Jellinger & Attems, 2015). A number of chronic health conditions have been shown to be significant independent predictors of ADRD (such as hypertension, diabetes, midlife obesity, cardiovascular and cerebrovascular disease (Baumgart et al., 2015) which may not only accelerate decline in demented elders (Melis et al., 2013), but aggravate functional impairment (Calderón-Larrañaga et al., 2019) and age-related, non-ADRD neurodegeneration, subsequently progressing into pronounced ischemic neurotoxicity, neuroinflammation, and other ADRD-related proteinopathies (Duthie et al., 2011); such neurologic pathologies have been documented in chronic pain sufferers as well (Cao, Fisher, Yu, & Dong, 2019). A growing body of research implicates the presence of chronic pain in neurobiological and neurochemical imbalances, persistent cellular stress and elevated pro-inflammatory mediators – pathological processes involved in cognitive deterioration and development of ADRD-related diseases (Moriarty, McGuire, & Finn, 2011). Collectively, these findings support a likely contribution of multimorbidity and chronic pain in promoting cognitive dysfunction and neuropathologic changes, and, together with the probable reciprocal relationship between chronic pain and multimorbidity, suggest that these conditions may have adverse synergistic effects on ADRD risk.

In this study, we found that the joint effects of NCPC’s and multimorbidity on ADRD risk were attenuated in strength and magnitude after inclusion of depression, anxiety, and insomnia-related sleep disorders, suggesting that these conditions may partially mediate the observed effects of co-occurring NCPC and multimorbidity on risk for incident ADRD. Multimorbidity, like NCPCs, has also been strongly and reciprocally associated with mental health conditions such as depression (Read et al., 2017), as well as insomnia-related sleep disorders (Helbig et al., 2017), factors directly linked to elevated risk for cognitive decline and ADRD. Presence of multimorbidity and NCPCs, through their role in worsening mental health (Read et al., 2017; Zis et al., 2017), can further exacerbate existing chronic health conditions, resulting in a vicious cycle of declining physical and mental health, increasing burden of chronic pain, and deterioration of cognitive function. While it can be argued that analgesic medications may in part explain the observed associations of NCPCs to ADRD risk, findings to date regarding the effects of opioid and NSAIDs analgesics use on cognitive function have been inconsistent (Etminan et al., 2003; Kurita, Pimenta, & Nobre, 2008). In this study, the use of neither NSAIDs nor opioid analgesics was related to risk for ADRD, nor did adjustment for these factors affect the joint or interactive effects of NCPC and multimorbidity on ADRD risk.

### Limitations

One of the major limitations of this study was the shorter duration of follow-up. As we know of ADRD being an underdiagnosed condition with ambiguous manifestations, the shorter follow-up duration may have led to an under-ascertainment of ADRD, pulling the risk toward the null. Moreover, claims data were used to ascertain multimorbidity status, which may suffer from under-reporting, potentially diluting the effect of multimorbidity in the analysis. Furthermore, the measure of number of chronic conditions, implying high load of multimorbidity, does not account for the type and different combinations of chronic health conditions constituting multimorbidity load, and that certain combinations contributing to multimorbidity may carry more weight in terms of risk factors for ADRD than others. Therefore, multimorbidity categories (2-3 and ≥ 4 chronic conditions) may not always reflect relative disease burden. Since we desired to assess the contribution of NCPCs and multimorbidity, first separately and then in tandem, it is important to note that our construct of multimorbidity only included select chronic physical health conditions while excluding any non-cancer chronic pain conditions as well as potential effect mediators including mental- and insomnia-related disorders. Additionally, the information on severity and stage of baseline chronic conditions was not available, and hence could not be included in our assessment of multimorbidity. Similarly, insufficient information on the severity or duration of NCPCs or chronic health conditions at the time of ascertainment hindered an in-depth assessment of the potentially dynamic influence of multimorbidity on the association of NCPCs to ADRD risk. Moreover, ADRD, multimorbidity and chronic pain have been associated with increased mortality, thereby, introducing potential survivor bias through overrepresentation of low-risk individuals (alive at the end of study follow up) in our analyses.

## 5. Conclusion

In this large population-based study of older Medicare beneficiaries, co-occurring NCPC and multimorbidity were associated with significantly elevated risk for ADRD, with ADRD risk highest in those with the greatest number of chronic conditions. If confirmed in future longitudinal investigations with longer follow-up durations and sophisticated measures of multimorbidity burden, these findings may aid in targeting at risk populations, help further motivate interventions designed to reduce NCPC and multimorbidity, and ultimately, inform strategies for decreasing ADRD risk. These findings also argue that the potential modifying effects of multimorbidity should be considered when assessing the effects of chronic pain on risk for cognitive impairment and ADRD.

## Data Availability

https://www.cms.gov/Research-Statistics-Data-and-Systems/Research-Statistics-Data-and-Systems

## Author contributions

S.K., K.E.I., U.S., contributed to research conceptualization, design, and methodology, and to the interpretation and presentation of results. S.K. and U.S. conducted the statistical analyses. S.K. prepared the manuscript draft; S.K., K.E.I., and U.S. worked on subsequent iterations. K.E.I., U.S., A.U., C.L., D.G., provided critical review of the final draft. All authors have read and agreed to the published version of the manuscript.

## IRB Statement

The present study was determined by the WVU Institutional Review Board (IRB) as not meeting the definition of human subject research due to the deidentified nature of the data used in our analyses (H-22407).

## Supplementary Material

Supplementary Table 1 Baseline characteristics of study participants by incident Alzheimer’s disease and related dementias (ADRD), MCBS and FFS claims data, 2001– 2013.

